# Regional differences in buprenorphine distribution in Pennsylvania from 2010-2020

**DOI:** 10.1101/2022.07.01.22277139

**Authors:** Alden J. Mileto, Robert J. Rinaldi, Steven J. Grampp, Kenneth L. McCall, Brian J. Piper

## Abstract

**Background:** Buprenorphine is a synthetic opioid frequently used in combination with naloxone for the treatment of opioid use disorder (OUD). Overall buprenorphine distribution has increased nationally; however, pronounced regional differences in this distribution have also been identified. The objective of this study was to analyze buprenorphine distribution by three-digit zip codes in Pennsylvania from 2010-2020.

**Methods:** Data was extracted from the Drug Enforcement Administration’s Automated Reports and Consolidated Orders System (ARCOS) yearly to gather buprenorphine distribution, in grams per 3-digit zip code, from 2010-2020. After compiling this data, a percent change for each 3-digit zip code was calculated to analyze the change in distribution from 2010-2020. The total weight of buprenorphine distributed for the state of Pennsylvania over the last decade was calculated. The amount of buprenorphine distributed in grams per each 3-digit zip code was compared to their population densities to analyze if there was any association between population and buprenorphine distribution. Zip codes that were outside of a 95% confidence interval were considered significant (*p* < .05).

**Results:** Pennsylvania pharmacies and hospitals dispensed 116.3 kg of buprenorphine in 2010. That number increased 217.3% to 369.0 kg in 2020. The 155-zip code (Somerset) experienced the largest increase (885%). In contrast the 190-zip (Philadelphia) experienced the smallest increase (79%). The 155 (Somerset), 169 (Wellsboro), and 177 (Williamsport) zip codes experienced significantly greater elevations relative to the state average.

**Conclusion:** Our analysis uncovered that buprenorphine distribution in Pennsylvania from 2010-2020 rose 217%. With the increasing awareness of opioid addiction, and the large number of opioid prescriptions in the US, this increase was expected. The zip codes of 155, 169, 177 showed a statistically significant increase in buprenorphine distribution relative to the overall state average. No zip codes displayed a statistically significant decrease in buprenorphine distribution. Interestingly, some of the more densely populated areas in Pennsylvania were at or below the average state increase of 217% (Pittsburgh 150-152 – 228%; Philadelphia 190-191 – 79%; Harrisburg 170-171 – 202%). Furthermore, the statistically significant zip codes of 155, 169, and 177 were among the least densely populated areas of Pennsylvania. Further pharmacoepidemiological research is needed to continue to characterize, and ideally remediate, the pronounced regional variation in buprenorphine distribution.

## Introduction

During the expansion of analgesics in the 1980s and 1990s, physicians started to treat pain with different modalities. Several scientific studies at the time stated that people were unable to become addicted to opiates (1). These beliefs, along with the push from pharmaceutical companies with statements like “Not treating patient pain with opiates is inhumane”, among other factors, lead to widespread prescription of opiates (1). Data suggests that in 2015 alone, approximately 2.4 million people in America suffered from an opioid use disorder (OUD) (2). Pennsylvania is a substantial contributor to the extensive opioid use in the US. In 2016, 4,642 individuals died from an opioid-related overdose in Pennsylvania (3). The consequential rise in opioid misuse and addiction lead to a declaration of a national public health emergency in 2017 (4).

Buprenorphine is a partial opioid agonist used to treat acute and chronic pain but is also indicated for treating opioid dependence. It is a Schedule III narcotic, meaning it has moderate or low physical dependence or high psychological dependence in pain management (5,7). Buprenorphine works by partially agonizing the mu opioid receptor, leading to a less intense activation that is seen with full agonists, such as morphine (5,7). Buprenorphine also acts as a weak kappa receptor antagonist and delta receptor agonist (5,7). It can be given independently or concurrently with naloxone. The latter combination allows for use of a partial opioid, without the achievement of full opioid effects. The use of buprenorphine was associated with a 32% relative rate of reduction in serious opioid-related acute care use at 3 months and a 26% relative rate of reduction at 12 months compared with no treatment (6)

Historically, the drug was developed in the 1960s as an alternative to stronger, full opioid agonist pain medications like morphine (7). In 2002, the Food and Drug Administration approved the drug for use in treating opiate disorders (2). This led to a larger distribution of buprenorphine as a treatment for OUD. In 2003, 11% of Opiate Treatment Programs (OTP) and about 5% of non-Opiate Treatment Programs (non-OTP), in the US, offered buprenorphine as a treatment method. By 2015, those numbers rose to 58% and 21% respectively (8). During the COVID-19 pandemic, OUD patients could be initiated with buprenorphine, but not methadone, pharmacotherapy (9). Pennsylvania ranked tenth in the US in 2017 for per capita buprenorphine distribution (10).

The purpose of this study was to analyze the trends in buprenorphine distribution, overall and by three-digit zip codes, in Pennsylvania from 2010-2020.

## Methods

### Procedures

Data was obtained from the Drug Enforcement Administration’s Automated Reports and Consolidated Orders System (ARCOS) database. The ARCOS database reports on the amount of each drug distributed by different issuance modalities (i.e., pharmacies and hospitals) and has been used in prior research (10). Each three-digit zip code in Pennsylvania (e.g., Scranton = 185) is broken down into quarterly and yearly reports, reported in total grams of drug distributed. Procedures were approved by the institutional review board of Geisinger and the University of New England.

### Data-analysis

The programs GraphPad Prism and Microsoft Excel were used to graph and analyze data. In order to normalize the distribution and adjust for the various population densities, number of pharmacies, and differing distributions between each three-digit zip code, a percent change for each three digit-zip code was calculated. To account for less populated areas having fewer total grams of distribution, the ratios were normalized by comparing the zip codes to themselves. The percent change was calculated by comparing each individual zip code’s total grams of buprenorphine distributed in 2010 to their totals in 2020. A 95% confidence interval was then calculated to determine if there was a statistically significant change in buprenorphine distribution as expressed as mean ± (1.96 x SD).

To display the regional variation, the percentage changes for three-digit zip codes were displayed on a heat map. An add on to Excel, Someka, was used to plot data points onto geographical locations. The percent change of each three-digit zip code was applied to each five-digit zip code. For example, the percent change of zip code 150 was 291%. Therefore, 291% was input for every five-digit zip code starting with 150.

## Results

The year with the largest total buprenorphine distribution between 2010 and 2020 was 2020, with 369,000.54 grams (369.0 kg) distributed. Comparing this to the year in which buprenorphine distribution was the lowest, 2010, which had a total of 116,301.74 grams (116.3 kg), there was a 217.3% increase (Figure 1).

**Figure 1.**
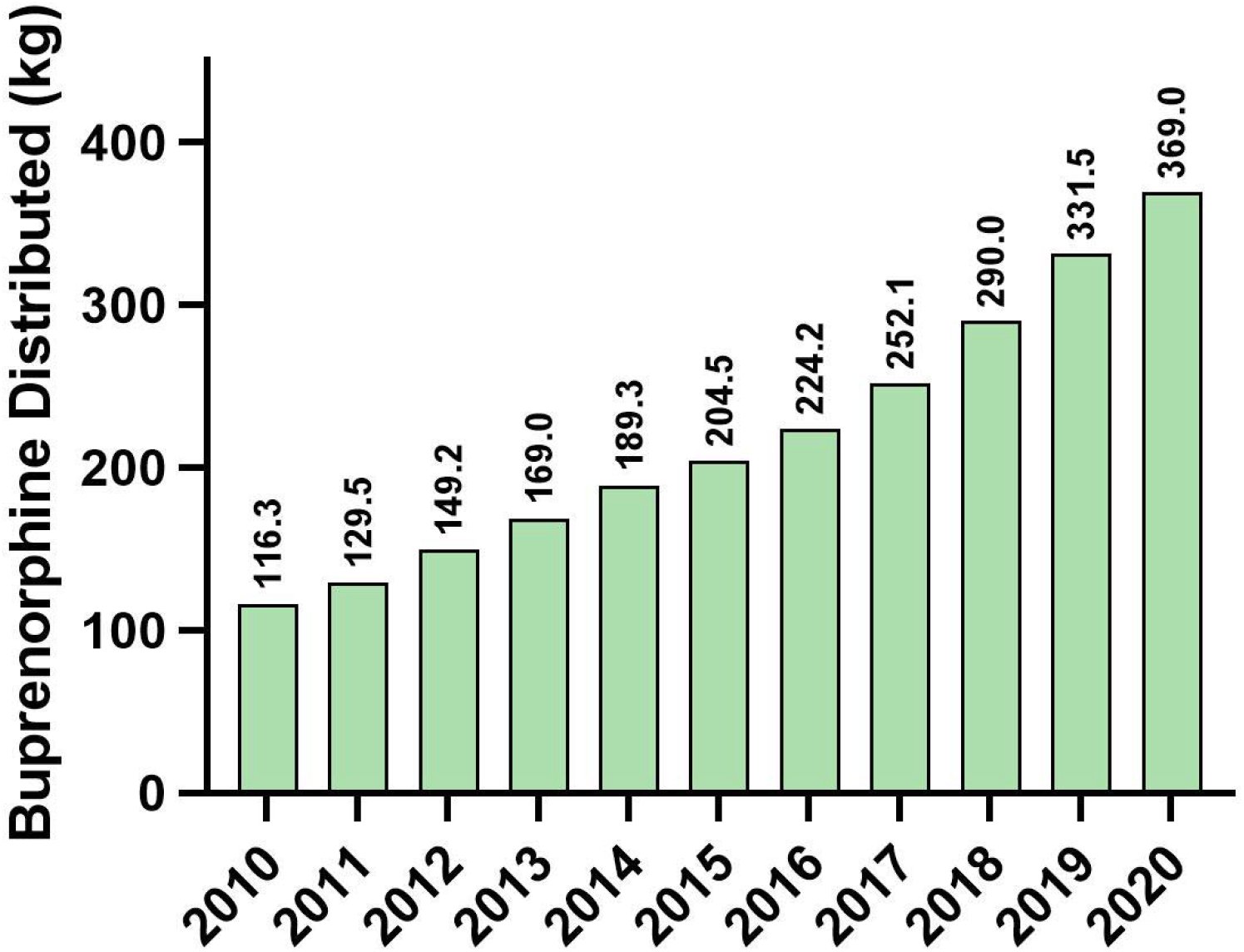
Buprenorphine distribution in total grams from 2010-2020 in Pennsylvania as reported by the Drug Enforcement Administration’s Automated Reports and Consolidated Ordering System (ARCOS). The percent change in buprenorphine distribution in Pennsylvania from 2010-2020 was 217%.

Total buprenorphine distribution was further broken down by 3-digit zip codes. There was a total of 47 three-digit zip codes. In 2010, the 190 (Philadelphia) zip-code had 15.2 kg of buprenorphine distributed. In 2020, the same zip code had 27.1 kg. The percent change in buprenorphine distributed was 79% which was the lowest percent change of all three-digit zip codes in the state (Figure 2). In 2010, the zip code 155 (Somerset) had 0.43 kg of buprenorphine distributed. In 2020, the same zip code had 4.2 kg. The percent change in buprenorphine distributed was 885% which was the highest of all three-digit zip codes (Figure 2).

**Figure 2.**
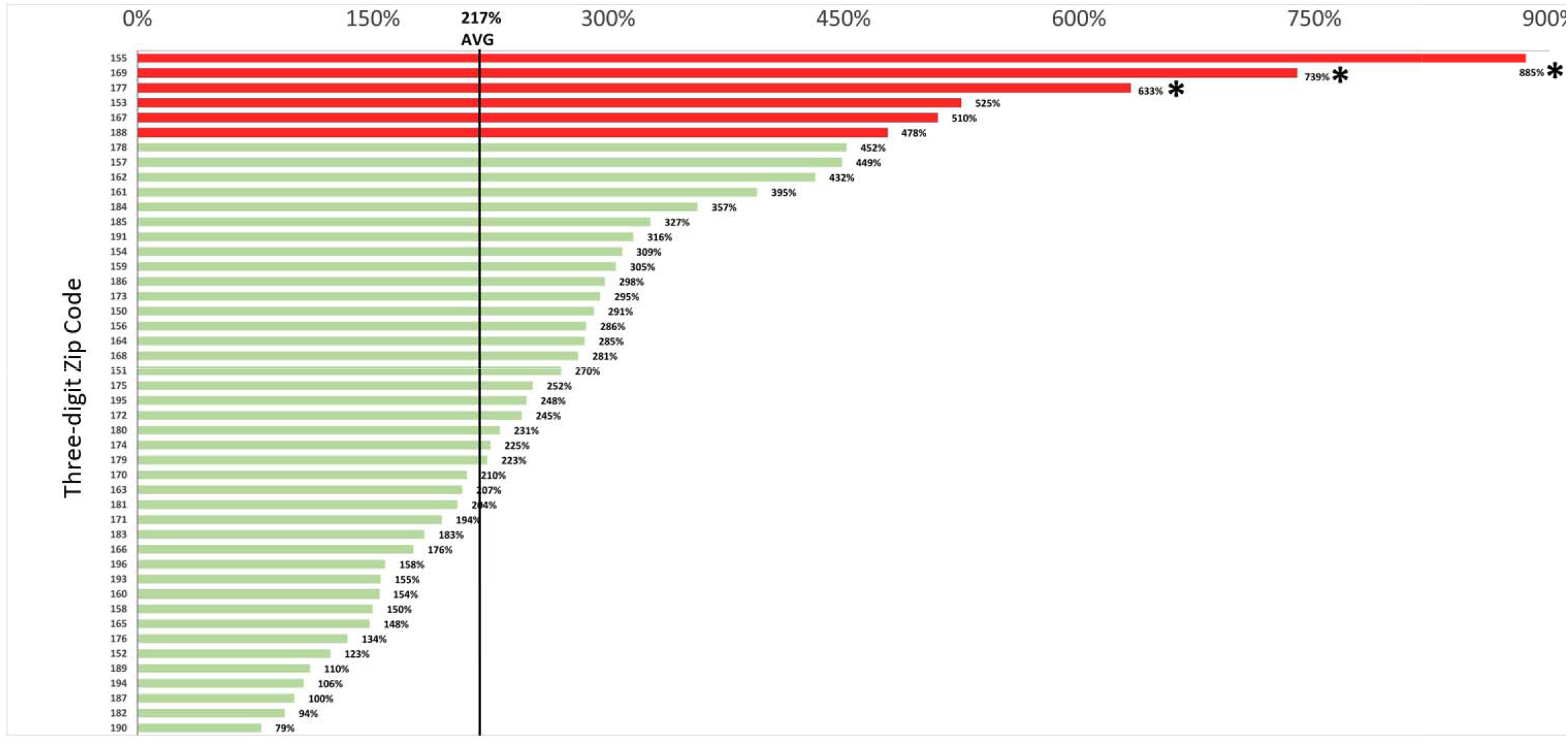
Percent change in buprenorphine distribution as reported by the Drug Enforcement Administration’s Automated Reports and Consolidated Ordering System (ARCOS) across three-digit zip codes in Pennsylvania from 2010-2020. The vertical line demarcates the average percent increase of 217% calculated from the total grams of buprenorphine distributed in PA from 2010-2020. Zip code values outside of a 95% confidence interval, calculated as mean ± (1.96 x standard deviation), were marked statistically significant with an asterisk. Zip codes +/-1.50 x standard deviation were marked in red.

No zip code showed a statistically significant decrease (i.e., outside of a 95% confidence interval) in the percent change in buprenorphine distribution. Zip codes 155 (Somerset), 169 (Wellsboro), and 177 (Williamsport) yielded statistically significant increases of 885%, 739%, and 633%, respectively (Figure 2). Figure 3 visualizes these regionally disparate patterns across the state. Refer to supplemental figure for reference of Pennsylvania population densities.

**Figure 3.**
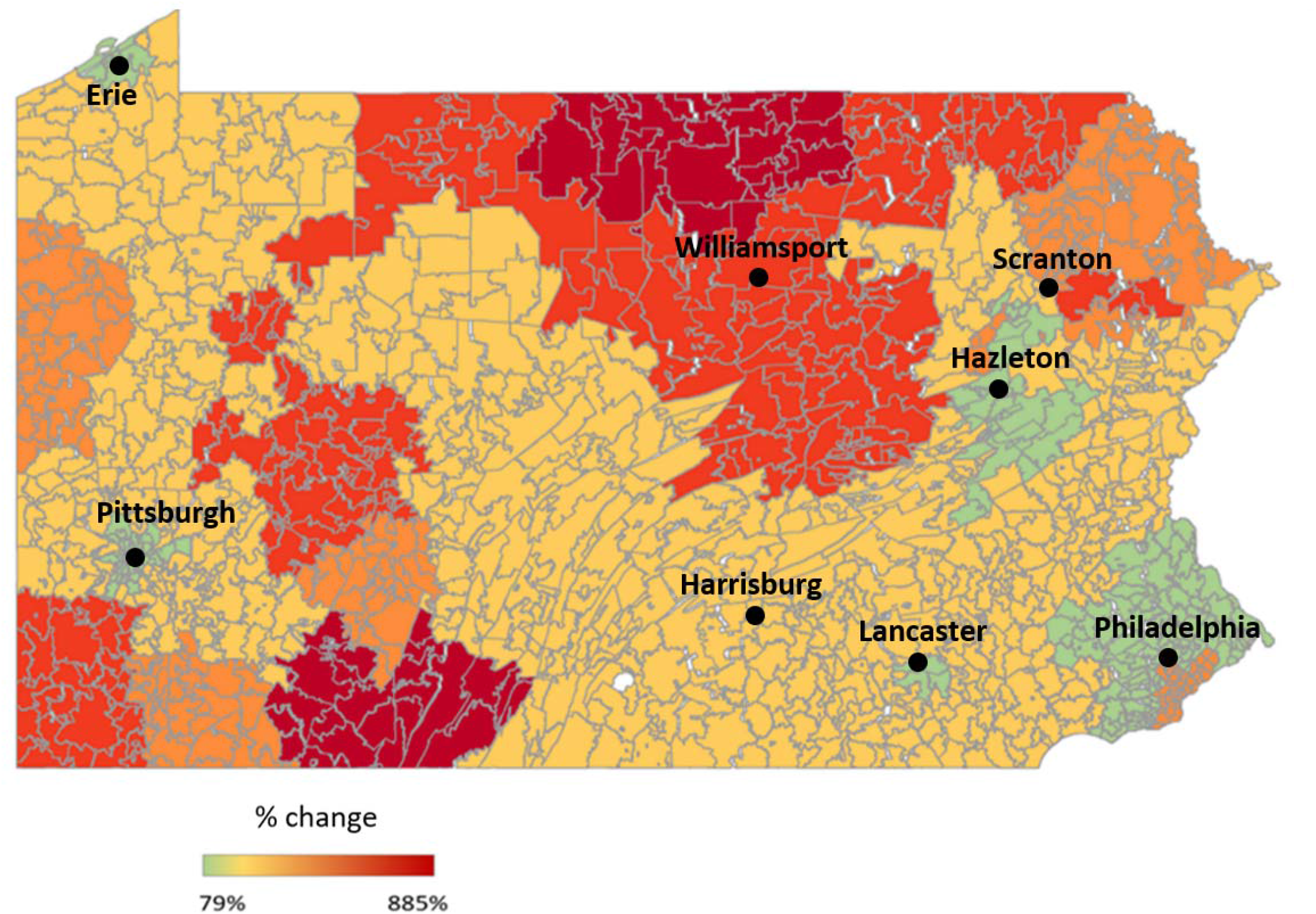
Percent change in buprenorphine distribution by zip codes as reported to the Drug Enforcement Administration’s Automated Reports and Consolidated Orders System (ARCOS) from 2010 to 2020.

## Discussion

The key finding of this report was that buprenorphine distribution in Pennsylvania increased from 2010 to 2020 by 217% (Figure 1). From 2009 to 2018, buprenorphine distribution across the United States rose by approximately 130% (8). From 2010 to 2018, buprenorphine distribution in Pennsylvania rose 149% (Figure 1). The rise in buprenorphine distribution in PA is comparable to the rest of the United States (8). However, studies show that buprenorphine use has been disproportionally distributed to rural or suburban areas in comparison to urban areas (10,12,15). This study and previous literature show a similar consensus (10).

Buprenorphine distribution in more densely populated areas of Pennsylvania were near or below the average of 217% (Pittsburgh: 150-152 – 228%; Philadelphia: 190-191 – 79%; Harrisburg: 170-171 – 202%, Figures 1 & 2). At the same time, the three-digit zip codes that had the highest percent increase in distribution belong to the three of the least densely populated areas (Somerset: 155 – 885%, Wellsboro: 169 – 739%, Williamsport: 177 – 633%) (Figures 1,2). These three-digit zip codes: 155, 169 and 177, had statistically significant increases relative to the rest of Pennsylvania. These statistics, showing a lack of buprenorphine distribution to densely populated areas of Pennsylvania, are troublesome given that Philadelphia and Allegheny County rank highest in rates of opioid related deaths amongst US counties with a population over 1 million people (13).

It was not an objective of this study to explore why the difference in percent change in buprenorphine distributions exists for each population density. However, some factors that may account for the data displayed are explored here. Although buprenorphine is effective in treating OUD (14), the lack of homogenous distribution is apparent (10,12,15). Studies show that roughly 56% of US counties that have the greatest need for buprenorphine treatment likely demonstrate inadequate measures to be able to use buprenorphine as an effective treatment (15). In Philadelphia, buprenorphine access disparities for minorities, in particular Hispanic populations and foreign-born, non-citizen populations, may exist because these populations have the highest documented uninsurance rates (16). The lack of resources in higher populated areas may play a role in the ability to prescribe buprenorphine.

Methadone is another evidence-based treatment for OUD. It has been demonstrated that buprenorphine and methadone are equally effective in treating OUD (17). However, buprenorphine is inferior to methadone in retaining patients in treatment (14). Studies show that in comparison to methadone, buprenorphine was more likely to be prescribed to white individuals (92% vs 53% of methadone patients), employed (56% vs 29% of methadone patients) and who have had some college education (56% vs 19% methadone) (12). White individuals make up approximately 80% of the rural population and 56% of the urban population (18). An explanation for this phenomenon may be the fact that methadone was approved for use by the FDA in 1972 and urban populations were the first to prescribe methadone (19). The observed geographical disparities could reflect greater availability of methadone from narcotic treatment programs that are typically located in more urban areas (20,21) whereas buprenorphine is available from primary care providers. In Philadelphia, it was demonstrated that areas with the lowest access to primary care had higher concentrations of non-Hispanic blacks and a lower median household income, compared to areas of higher access to primary care (22).

## Limitations

This novel pharmacoepidemiological study has some potential limitations. ARCOS does not distinguish between the buprenorphine which is distributed to treat OUD versus the presumably much smaller subset, including by veterinarians (23), that was subsequently used to treat pain. Similarly, distribution information does not illuminate how much reached the intended patients versus how much of this Schedule III drug was diverted to others (24). Although the ARCOS data was reported at the level of three-digit zip codes, some patients, particularly those in rural areas, may not reside in the same zip code as the pharmacy where the buprenorphine was distributed. In addition, the consolidation of five-digit zip codes in order to represent one three-digit zip code decreased the granularity of regional percentage changes. It is also unclear whether the areas with the largest percent changes in buprenorphine distribution, had particularly low distribution rates in 2010, resulting in a relatively large percentage increase or whether there was a truly a disproportionate increase in distribution in these areas compared to other areas. Future research could be completed at the county or even patient level using electronic medical records to further characterize the regional, rural/urban, racial/ethnic, and socioeconomic disparities in OUD treatment in the US.

## Conclusion

This analysis uncovered that from 2010 to 2020 the percent increase in buprenorphine prescription in the state of Pennsylvania was 217%. With the increasing awareness of opioid addiction and the over-prescription of opioids in the US, along with additional physician, nurse practitioner, and physician assistant training in buprenorphine treatment delivery, this percent increase was expected. The zip codes of 155 (Somerset), 169 (Wellsboro), 177 (Williamsport) showed a statistically significant increase in buprenorphine distribution. Interestingly, these zip codes are in some of the least densely populated areas in Pennsylvania. No zip codes displayed a statistically significant decrease in buprenorphine distribution. However, some of the more densely populated areas of Pennsylvania were near or below the average of 217% (Pittsburgh 150-152 – 228%; Philadelphia 190-191 – 79%; Harrisburg 170-171 – 202%). This pattern warrants further investigation into the gaps of care in buprenorphine distribution in higher population dense areas. Overall, this study provides a foundational basis for investigation of additional opioid and opioid treatment patterns in Pennsylvania and other states that continue to be adversely impacted by the iatrogenic US opioid epidemic.

## Data Availability

All data is available online at: https://www.deadiversion.usdoj.gov/arcos/retail_drug_summary/index.html

## Contributor Roles

Alden J. Mileto preformed literature search and review, analyzed the data, prepared figures, authored drafts of the paper, and approved the final manuscript.

Robert J. Rinaldi preformed literature search and review, analyzed the data, prepared figures, authored drafts of the paper, and approved the final manuscript.

Steven J. Gramp preformed literature search and review, analyzed the data, authored drafts of the paper, and approved the final manuscript.

Kenneth L. McCall provided feedback on data analysis and interpretation and approved the final manuscript.

Brian J. Piper authored drafts of the paper, analyzed data, and approved the final manuscript.

## Institutional Review Board Statement

This study was deemed exempt by the IRB of Geisinger 2020-0223.

## Acknowledgments

We would like to thank Poul Chinga, MBS for his guidance during the data analysis process. We would like to thank Olivia Lattanzi for her guidance and feedback during the editorial process. This study was supported by the Health Resources Services Administration (D34HP31025).

---

**Supplemental Figure 1.**
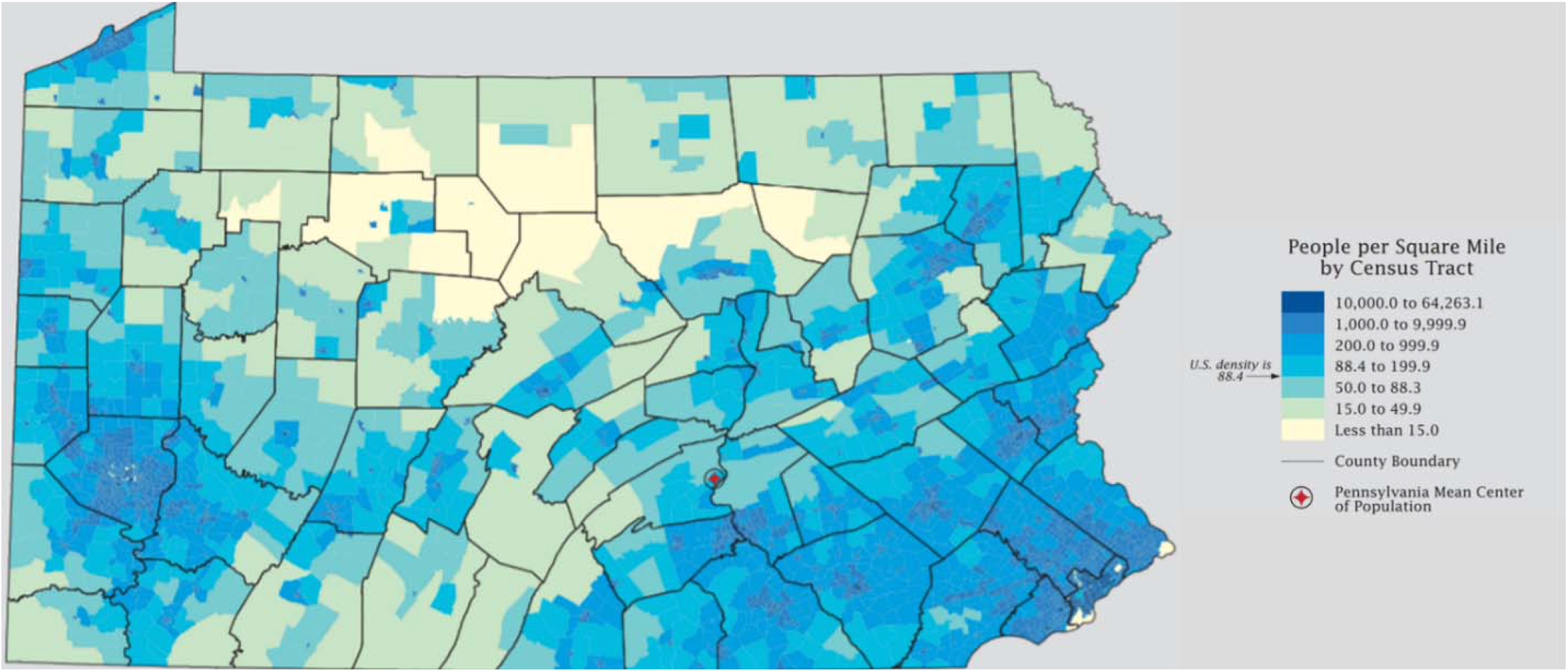
Population densities of Pennsylvania by zip code and county according to the 2010 Census: Pennsylvania Profile (11).

## References

1. Jones MR, Viswanath O, Peck J, Kaye AD, Gill JS, Simopoulos TT. A brief history of the opioid epidemic and strategies for pain medicine. Pain Ther. 2018;7(1):13–21. doi: 10.1007/s40122-018-0097-6

2. Alderks CE. Trends in the use of methadone, buprenorphine, and extended-release naltrexone at substance abuse treatment facilities: 2003-2015 (Update). The CBHSQ Report. Rockville (MD) 2013. p. 1-8. PMID: 29200242

3. U.S. Department of Health and Human Services. Geographic Disparities Affect Access to Buprenorphine Services for Opioid Use Disorder. Published 2020. Accessed March 10, 2022. https://oig.hhs.gov/oei/reports/oei-12-17-00240.pdf

4. Pennsylvania Office of Attorney General. Opioid Battle. Published 2022. Accessed March 14, 2022. https://www.attorneygeneral.gov/protect-yourself/opioid-battle/

5. Kumar E, Viswanath O, Saadabadi A. Buprenorphine. StatPearls. 2022. PMID: 29083570

6. Wakeman SE, Larochelle MR, Ameli O, Chaisson CE, McPheeters JT, Crown WH, et al. Comparative effectiveness of different treatment pathways for opioid use disorder. Jama Network Open. 2020;3(2). doi: 10.1001/jamanetworkopen.2019.20622

7. Pergolizzi J, Aloisi AM, Dahan A, Filitz J, Langford R, Likar R, et al. Current knowledge of buprenorphine and its unique pharmacological profile. Pain Pract. 2010;10(5):428–50. doi: 10.1111/j.1533-2500.2010.00378.x

8. Olfson M, Zhang V, Schoenbaum M, King M. Trends in buprenorphine treatment in the United States, 2009-2018. Jama-J Am Med Assoc. 2020;323(3):276–7. doi:10.1001/jama.2019.1891

9. SAMHSA. FAQs: Provision of methadone and buprenorphine for the treatment of opioid use disorder in the COVID-19 emergency. Rockville, MD: Substance Abuse and Mental Health Services Administration. Published 2020. Accessed March 20, 2022. https://www.samhsa.gov/sites/default/

10. Pashmineh AR, Cruz-Mullane A, Podd JC, Lam WS, Kaleem SH, et al. Rise and regional disparities in buprenorphine utilization in the United States. Pharmacoepidemiol Drug Saf. 2020;29(6):708–715. doi: 10.1002/pds.4984.

11. U.S. Department of Commerce Economics and Statistics Administration. 2010 Census: Pennsylvania Profile. Published 2010. Accessed March 10, 2022. https://www2.census.gov/geo/maps/dc10_thematic/2010_Profile/2010_Profile_Map_Pennsylvania.pdf

12. Hansen HB, Siegel CE, Case BG, Bertollo DN, DiRocco D, Galanter M. Variation in use of buprenorphine and methadone treatment by racial, ethnic, and income characteristics of residential social areas in New York City. J Behav Health Serv Res. 2013;40(3):367–77. doi: 10.1007/s11414-013-9341-3

13. Commonwealth Prevention Alliance. PAStop. Published 2022. Accessed March 10, 2022. https://pastop.org/about/

14. Mattick RP, Kimber J, Breen C, Davoli M. Buprenorphine maintenance versus placebo or methadone maintenance for opioid dependence. Cochrane Database Syst Rev. 2008;(2):CD002207. doi: 10.1002/14651858.CD002207.pub2

15. U.S. Department of Health and Human Services. Geographic Disparities Affect Access to Buprenorphine Services for Opioid Use Disorder. Published 2020. Accessed March 10, 2022. https://oig.hhs.gov/oei/reports/oei-12-17-00240.pdf

16. Philadelphia Department of Public Health. Access to primary care in Philadelphia. CHART 2019;4(8):1–5.

17. Wakeman SE, Larochelle MR, Ameli O, Chaisson CE, McPheeters JT, Crown WH, et al. Comparative effectiveness of different treatment pathways for opioid use disorder. Jama Network Open. 2020;3(2). doi: 10.1001/jamanetworkopen.2019.20622

18. United States Department of Agriculture. Rural America at a glance. Published 2018. Accessed March 10, 2022. https://www.ers.usda.gov/webdocs/publications/90556/eib-200.pdf

19. Samet JH, Botticelli M, Bharel M. Methadone in primary care - one small step for congress, one giant leap for addiction treatment. New Engl J Med. 2018;379(1):7–8. doi: 10.1056/NEJMp1803982

20. Hyde TF, Bekoe-Tabiri AD, Kropp Lopez AK, Devia LG, Gutierrez BD, et al. County and demographic differences in drug arrests and controlled substance use in Maine. Journal of Maine Medical Center. 2021;3(2). doi: 10.46804/2641-2225.1074.

21. Furst JA, Mynarski NJ, McCall KL, Piper BJ. Pronounced regional disparities in United States methadone distribution. Ann Pharmacother. 2022;56(3):271–279. doi: 10.1177/10600280211028262.

22. Philadelphia Department of Public Health. Staying Healthy: Access to Primary Care in Philadelphia. 2018. Accessed June 24, 2022. https://www.phila.gov/media/20181109113640/2018-PrimaryCareReportFINAL.pdf

23. Piper BJ, McCall KL, Kogan LR, Hellyer P. Assessment of controlled substance distribution to U.S. veterinary teaching institutions from 2006 to 2019. Front Vet Sci. 2020; 7:615646. doi: 10.3389/fvets.2020.615646.

24. Hswen Y, Zhang A, Brownstein JS. Leveraging black-market street buprenorphine pricing to increase capacity to treat opioid addiction, 2010-2018. Prev Med. 2020;137:106105. doi: 10.1016/j.ypmed.2020.106105.

